# Machine-learning-aided multiplexed nanobiosensor for COVID-19 population immunity profiling

**DOI:** 10.1101/2023.02.06.23285535

**Authors:** Aidana Beisenova, Wihan Adi, S. Janna Bashar, Monniiesh Velmurugan, Kenzie B. Germanson, Miriam A. Shelef, Filiz Yesilkoy

## Abstract

Serological population surveillance can elucidate immunity landscapes against SARS-CoV-2 variants and are critical in monitoring infectious disease spread, evolution, and outbreak risks. However, current serological tests fall short of capturing complex humoral immune responses from different communities. Here, we report a machine-learning (ML)-aided nanobiosensor that can simultaneously quantify antibodies against the ancestral strain and Omicron variants of SARS-CoV-2 with epitope resolution. Our approach is based on a multiplexed, rapid, and label-free nanoplasmonic biosensor, which can detect past infection and vaccination status and is sensitive to SARS-CoV-2 variants. After training an ML model with antigen-specific antibody datasets from four COVID-19 immunity groups (naïve, convalescent, vaccinated, and convalescent-vaccinated), we tested our approach on 100 blind blood samples collected in Dane County, WI. Our results are consistent with public epidemiological data, demonstrating that our user-friendly and field-deployable nanobiosensor can capture community-representative public health trends and help manage COVID-19 and future outbreaks.

## Introduction

The COVID-19 pandemic has disrupted lives globally in various degrees since the outbreak of SARS-CoV-2 in 2019. While the worst has been abated in many parts of the world through vaccination efforts or various public health policies, a return to a COVID-19-free world is not expected, and the risk of new viral epidemics and pandemics is real ^[1]^, highlighting the need for new pandemic management strategies. Effective disease surveillance is a prerequisite for tenacious pandemic management through prompt implementations of public health measures. A major public health strategy is to ensure that the population is well protected against the virus and its emerging variants of concern (VOCs). Protective immunity against symptomatic infection with SARS-CoV-2 is well correlated with the presence of neutralizing antibodies ^[2,3]^. Thus, an effective approach to epidemiological immunity surveillance is anti-SARS-CoV-2 antibody monitoring at the population level ^[4,5]^. However, a comprehensive understanding of the clinically ^[6–8]^and demographically ^[9,10]^heterogeneous immune response is not straightforward, requiring antibody landscaping against multiple antigens and monitoring temporal dynamics and duration of antibody responses ^[11–13]^. Given variations in individuals ‘ humoral immunity as well as the rapid emergence of SARS-CoV-2 VOCs, serologic assays that can provide detailed information about protective immunity are needed for rigorous epidemiological surveys. Hence, serologic tests capable of antigen-specific multiplexed antibody quantification are paramount to immunity profiling against multiple VOCs of SARS-CoV-2 after infection and vaccination among diverse populations.

Some of the conventional serologic assays for SARS-CoV-2 are chemiluminescent immunoassays (CLIA) ^[7,14]^, enzyme-linked immunosorbent assays (ELISA) ^[15,16]^, and lateral flow assays (LFA) ^[16,17]^. CLIA and ELISA are widely used laboratory-based methods; however, they require multiple reagents and assay steps, non-portable optical readers, and trained operators, which impede their widespread and equitable access to diverse global populations. LFAs, on the other hand, are simple and cost-effective, yet they lack quantitative and multiplexed detection capabilities and suffer from performance variations ^[18]^. Notably, false-positive results are common in serologic assays based on a single antigen because some sera contain cross-reactive antibodies ^[19,20]^. Consequently, new optical biosensors are emerging as promising alternatives to gold-standard methods with quantitative and multiplexed antibody detection capabilities ^[21–26]^. However, they usually require exogenous labels, such as fluorophores, which suffer from photostability ^[27]^ and background interference ^[28]^. Label-free and reagent-less analyte quantification from small blood quantities is critical to developing simple bioassays and portable and low-cost optical readers that can be deployed for widespread immunity screening.

In addition to the need for simple assays, multiple clinical studies have demonstrated that antibody responses to specific viral epitopes correlate with COVID-19 clinical severity and patient survival ^[29,30]^. This finding highlights the importance of epitope-resolved antibody screening to elucidate the complex humoral immunity to SARS-CoV-2 and identify patient-specific neutralizing antibody patterns, which can serve as complex biomarkers and carry diagnostic and prognostic information. Moreover, the antibody response to different mutant peptides found in viral epitopes can elucidate the humoral immune response to SARS-CoV-2 VOCs. Yet, epitope-resolved multiplexed antibody screening tests are far from accessible, despite their important role in biomedical research, clinical diagnostics, and epidemiological applications. Additionally, high throughput serologic assays generate complex datasets that require advanced classifiers since the data cannot simply be interpreted using conventional cutoff thresholding. This complexity introduces a unique opportunity to leverage advances in machine learning (ML) by training serodiagnostic algorithms using high-dimensional epitope-resolved antibody datasets that are accurately measured by user-friendly and field-deployable biosensors.

Here, we introduce a multiplexed and label-free nanoplasmonic biosensor empowered by an ML model capable of discerning past infection and vaccination status that is sensitive to SARS-CoV-2 VOCs. Our nanoengineered photonic sensor chips are functionalized with six different SARS-CoV-2 antigens from ancestral strain (wild type, WT) and Omicron variants, including nucleocapsid (N) and spike (S) proteins, S protein ‘s receptor binding domain (RBD), and epitope-containing peptide from the membrane (M) protein, as well as two non-specific control proteins in spatially encoded microarrays. Our optical interrogation approach is based on widefield imaging; thus, we can simultaneously screen multiple antigen-specific antibodies from each immunoreaction microspot in a single assay step. The molecular detection mechanism is based on the nano-biosensor ‘s optical resonance shift upon antibody binding to antigen-immobilized sensor areas. Consequently, the sensor ‘s resonance response generates an image intensity contrast, which is directly correlated to antibody quantities in sera without the need for labeling steps (Fig 1b). Using our nano-biosensor platform, we measured sera from four sample groups with clinically known COVID-19 immune profiles: COVID-19 naïve, vaccinated without past infection (V), convalescent unvaccinated (C), and convalescent vaccinated (CV). After training an ML model (Random Forest) using these clinically labeled data, we tested the ML model by measuring and classifying 100 blind samples from Dane County, WI, USA, with unknown COVID-19 immune profiles (Fig.1). Our ML-aided nanobiosensor predicted 55% past infection (27% by the Omicron variant) and 87% vaccination rate based on the antibody data from blind samples. Notably, our findings are in good agreement with publicly available epidemiological data (65% past infection, in which 30% is Omicron, and 84% vaccination rate in Dane County, WI) at the time when our samples were collected. Our findings reveal the potential impact of our biosensor platform for widespread population health screening and pandemic management.

**Fig. 1.**
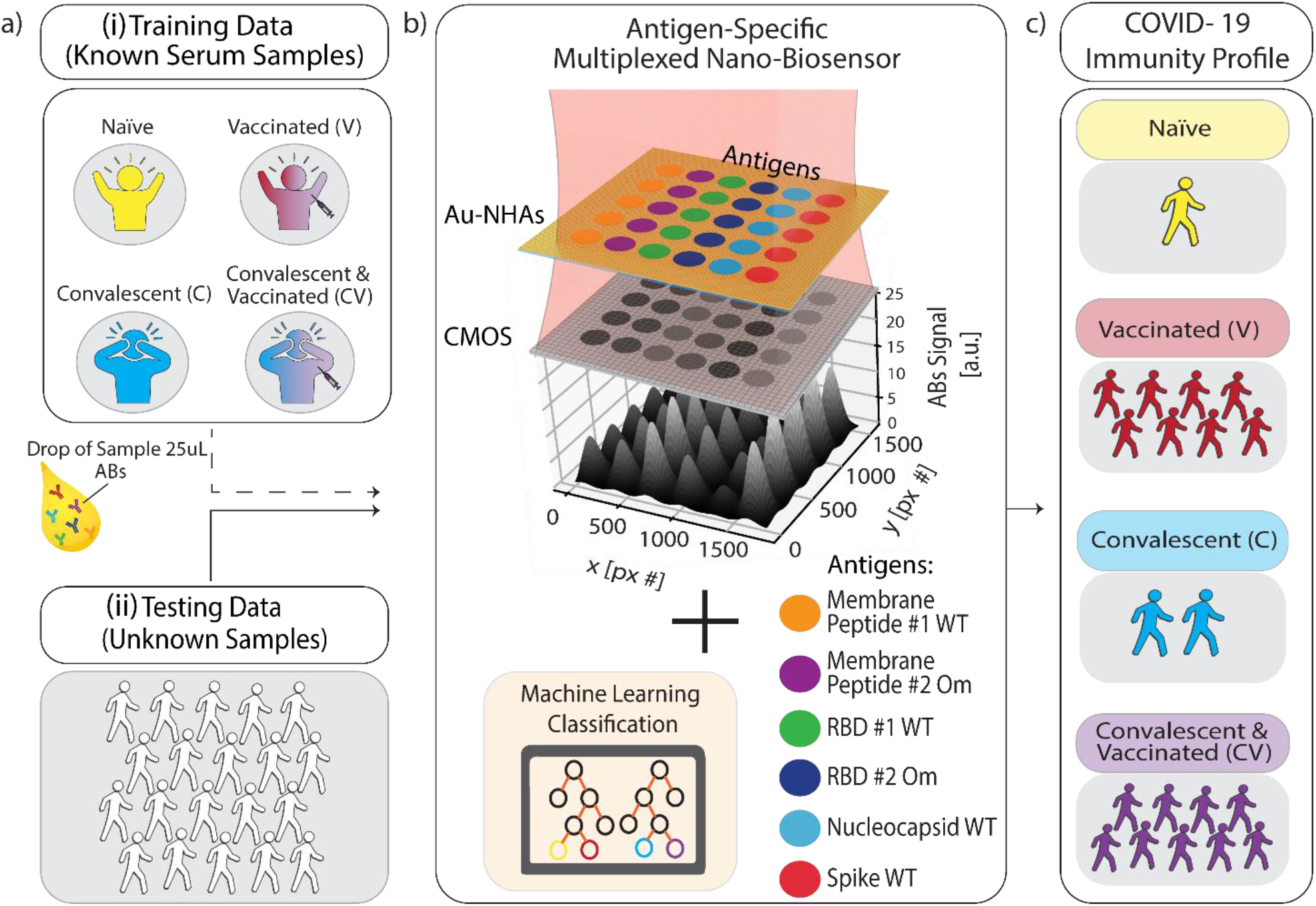
Schematic overview of multiplexed nanobiosensor technology coupled with an ML-based classification model to profile population-level immunity status against SARS-CoV-2. **(a)** Human serum or plasma samples (25 μl) from two groups were used: (i) known COVID-19 immunization and infection status (no past SARS-CoV-2 infection or vaccination (naïve), past COVID-19 (convalescent), and vaccinated with and without past SARS-CoV-2 infection) for ML training data, and (ii) blind samples for testing our approach. **(b)** Nanobiosensors were functionalized with six different antigenic proteins and peptides from SARS-CoV-2 in a microarray. Samples were loaded on sensor chips and measured by capturing a single image in bright-field transmission mode using a CMOS camera and single-wavelength illumination at 650 nm. Antigen-specific antibody binding on the biofunctionalized nanobiosensor surface generates image intensity contrast, which correlates to antibody (Abs) concentration in the sample. **(c)** An ML model classified each sample ‘s measured quantitative antibody datasets to obtain their COVID-19 immunity profiles.

## Results

### Antibody screening against SARS-CoV-2 antigens from clinically labeled samples

We performed antibody screening bioassays against SARS-CoV-2 antigens using plasmonic gold nanohole array sensors, which we recently reported in ^[26]^, and here we call them nanobiosensors. The antigens of interest are immobilized on the sensor surface using a noncontact liquid dispenser in a microarray format. The nanobiosensors were optically interrogated on a simple widefield imaging platform in transmission mode. The CMOS-camera-acquired microarray images were then used for multiplexed antibody quantification from human serum and plasma samples. We first collected reference images by illuminating the sensors with red light (λ=650 nm). Subsequently, we loaded samples (25 μl) into the measurement chamber, incubated them for 30 mins, washed them, and collected detection images. When the target antibodies are present in the samples, they specifically bind to the surface-bound antigens, decreasing the total transmittance intensity in the corresponding microarray spots. To measure antibody concentrations, we calculated the intensity contrast differences from the sensor images acquired before and after sample incubation for each antigen spot and correlated them with the antibody concentrations (illustrated in Fig. 1(b), for more details, see Materials and Methods). Finally, nanobiosensor performance was validated previously and achieved high concordance with standard CLIA (*26*).

To measure the immune response to SARS-CoV-2, we compared antibody levels against six SARS-CoV-2 antigens (see Fig.2) from the following groups of subjects: i) COVID-19 naïve collected before the emergence of SARS-CoV-2 as a control group, ii) vaccinated with no known past COVID-19 infection, iii) convalescent (i.e., positive past COVID-19 infection), and iv) convalescent and vaccinated. Each group contains 18 serum samples from different individuals, and only 25 μl sample volume was used for each measurement. Our reported antibody concentration values correspond to the total antibody response of all immunoglobulin isotypes and subclasses specific to the same antigen.

Fig. 3 shows the measured antigen-specific antibody levels of individuals from the four COVID-19 immunity groups. In COVID-19 naïve group (Fig. 3(a)), the average measured antibody values are negative across all antibodies, implying that no specific antibody binding to the viral antigens was detected. The only exception is positive antibody binding for anti-N antibodies (average signal 4.04 a.u.), likely due to cross-reactivity with common cold coronavirus N protein, which was also observed in a previous study ^[31]^. Otherwise, the low reactivity signals against all tested SARS-CoV-2 antigens are expected because the samples were collected before the onset of the pandemic.

For the vaccinated group (Fig. 3(b)), the average antibody levels against RBD and S protein are higher than the naïve controls (anti-RBD_WT p < 5 × 10^−7^ and anti-S_WT p < 5 × 10^−10^). We also observed that the measured concentration of anti-S_WT (average signal 26.39 a.u.) is distinctly higher compared to anti-RBD_WT (average signal 14.57 a.u., p < 5 × 10^−2^). This result is expected because the two widely applied mRNA vaccines (BioNTech-Pfizer and Moderna) contain sequences for full-length S protein ^[32]^. Thus, RBD, a smaller S protein fragment, elicits a lower total concentration of antibodies than the S protein.

Moreover, the antibody concentrations against RBD Omicron (anti-RBD_Om) were higher in the vaccinated group compared to the naïve group (p < 5 × 10^−5^) but not as high as anti-RBD_WT (p < 5 × 10^−7^). This finding implies that vaccination is more effective against the WT virus than the Omicron variant, consistent with the fact that all authorized vaccines in the US prior to sample collection were designed based on WT SARS-CoV-2^[33–35]^. The low antibody response to RBD_Om among vaccinated individuals can also be explained by the lack of past Omicron infections in this group and the relatively high number of mutations in the RBD of the Omicron variant, hindering the binding affinity of vaccine-induced anti-RBD antibodies (Fig. 2). Finally, the antibody values against M-peptides in the vaccinated group is the same as in naïve subjects due to the lack of past infections in this group and the fact that the vaccines do not contain M-protein coding mRNA sequences. Consistent with earlier findings ^[31]^, these data suggest that the M peptide (with sequence from ^[36]^) can be used as a biomarker of past infection. Finally, anti-N_WT antibody levels in both naïve and vaccinated groups are similar, with averages close to zero (naïve 4.04 a.u. and V 1.82 a.u.) and large standard deviations (naïve 14.20 a.u. and V 11.16 a.u.), again likely due to cross-reactivity with other coronaviruses.

**Fig. 2.**
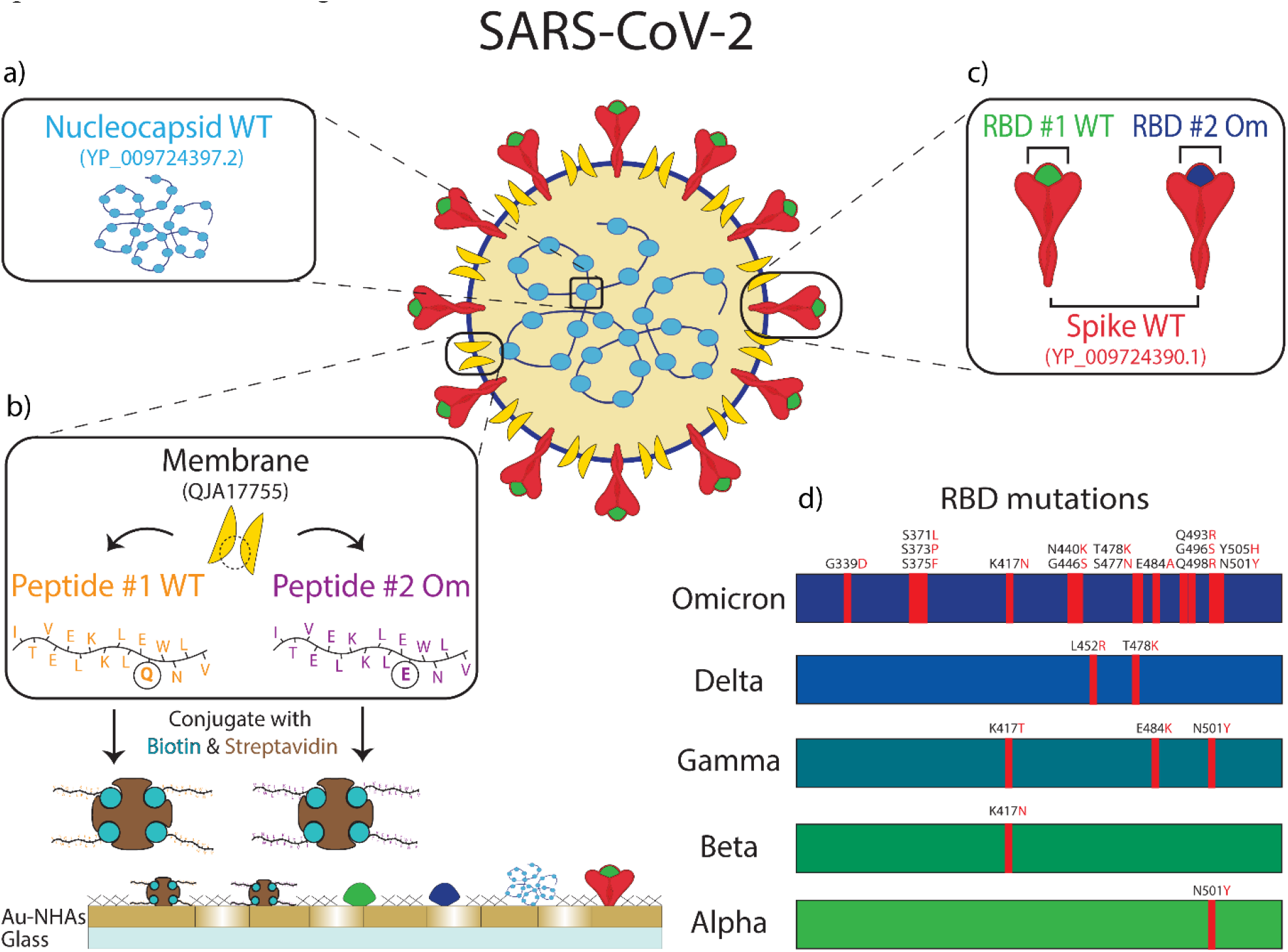
Six different antigenic proteins, protein domains, and peptides from SARS-CoV-2 are immobilized on the nanobiosensor surface: **(a)** Nucleocapsid protein from wild-type (WT) SARS-CoV-2. **(b)** Membrane peptide containing epitope from WT SARS-CoV-2 (Peptide #1 WT) and Omicron variant of SARS-CoV-2 (Peptide #2 Om). These biotin-conjugated membrane peptides were bound to streptavidin before immobilizing them on the sensor surface. **(c)** Spike protein from WT SARS-CoV-2, spike protein receptor binding domain (RBD) from WT SARS-CoV-2 (RBD #1 WT), and the Omicron variant of SARS-CoV-2 (RBD #2 Om). **(d)** The mutations in RBD for different variants are shown on the bar charts. The Omicron variant with 15 mutations is compared to the WT and other early variants for reference.

In the convalescent group (Fig. 3(c)), samples were collected in the spring of 2020, ∼5 to ∼12 weeks after infection onset. Here, infection-elicited antibodies against all SARS-CoV-2 antigens are significantly higher than naïve group (anti-M_WT p < 5 × 10^−10^, anti-M_Om p < 5 × 10^−9^, anti-RBD_WT p < 5 × 10^−10^, anti-RBD_Om p < 5 × 10^−3^, anti-S_WT p < 5 × 10^−10^, anti-N_WT p < 5 × 10^−6^). Moreover, in the convalescent group, anti-RBD_WT levels are higher than anti-RBD_Om (Fig. 3(c), p < 5 × 10^−6^), which is expected because the samples are from before the emergence of the Omicron variant. Similar to the vaccinated group, anti-S_WT levels are consistently higher than anti-RBD antibody levels in the convalescent group. Moreover, infection-induced anti-M peptide antibody responses are significantly higher than in naïve and vaccinated groups. This observation is consistent with previous studies ^[31,36]^ and underscores that seropositivity to M-protein peptides can help differentiate convalescent from vaccinated subjects. The slightly lower average antibody recognition difference for M_Om compared to M_WT peptide is not statistically significant. The variant sensitivity in antibody recognition for RBD compared to M-protein peptide may be due to the higher number of mutations in RBD (15 vs. 1 amino acid difference) than the M-protein.

**Fig. 3.**
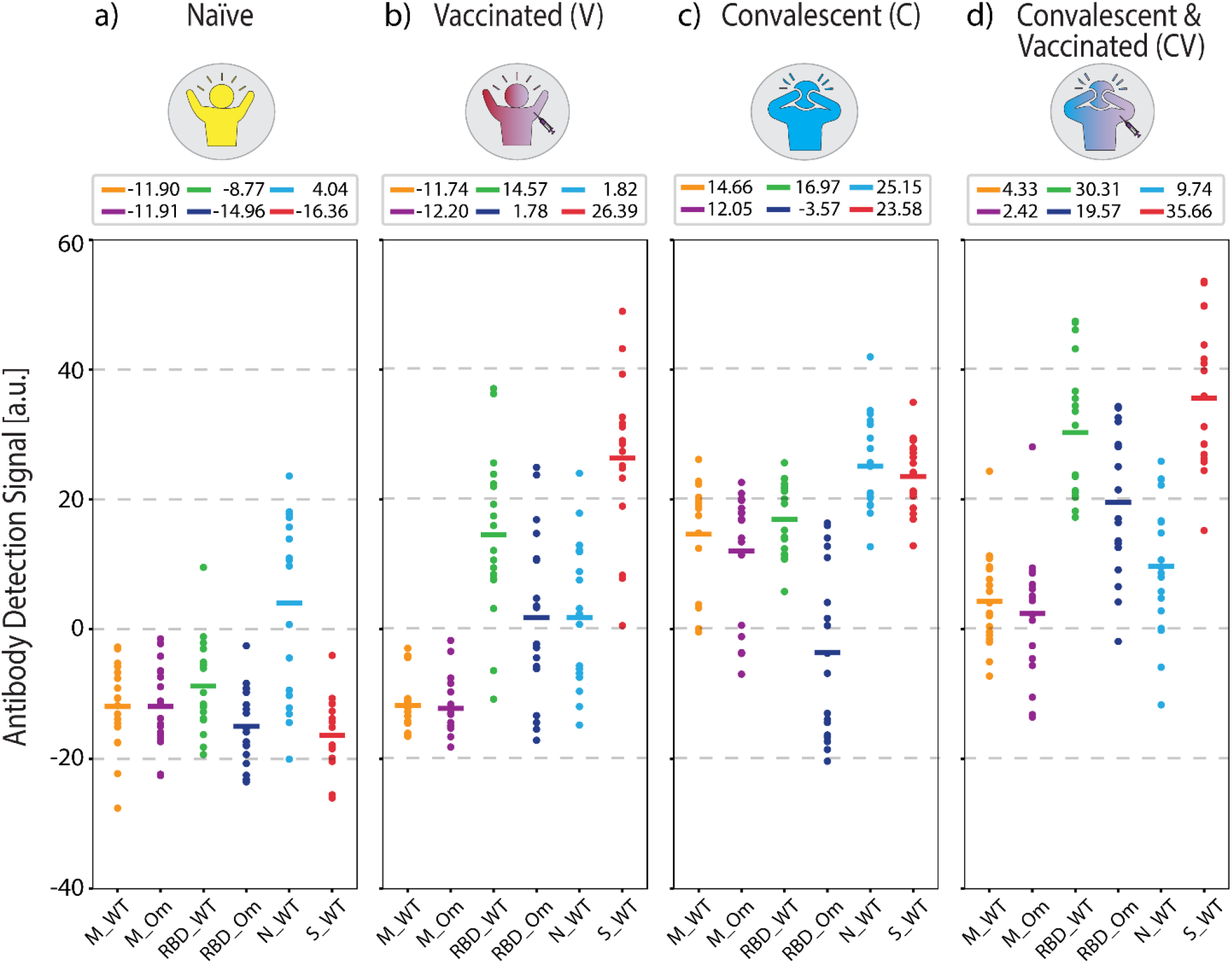
Antigen-specific antibody measurements from samples with known COVID-19 status used in ML model training. **(a)** COVID-19-naïve, **(b)** vaccinated, **(c)** convalescent, and **(d)** convalescent and vaccinated group antibody levels measured from n=18 individuals per group. Solid lines indicate mean values, which are also written above each plot. Dashed lines are for visual guidance.

Finally, we quantified antibody responses in the convalescent-vaccinated samples (Fig. 3(d)). All three S-protein antigens showed a further increase in reactivity compared to the only vaccinated group (anti-RBD_WT p < 5 × 10^−5^, anti-RBD_Om p < 5 × 10^−5^, anti-S_WT p < 5 × 10^−2^) and an increase compared to the convalescent group (anti-RBD_WT p < 5 × 10^−5^, anti-RBD_Om p < 5 × 10^−6^, anti-S_WT p < 5 × 10^−5^). Notably, the difference in antibodies against RBD between the two tested variants is smaller for the convalescent-vaccinated group than convalescent group (RBD_WT 16.97 a.u. and RBD_Om -3.57 a.u., p < 5 × 10^−6^ for the convalescent group vs. RBD_WT 30.31 a.u. and RBD_Om 19.57 a.u., p < 5 × 10^−2^ for the convalescent-vaccinated group). Given that the samples were collected before the onset of the Omicron variant, the increase in RBD_Om could be attributed to an overall increase of anti-S antibodies or a broader immune response after the multiple SARS-CoV-2 S protein exposures. Like in the vaccinated group, the vaccine boosted the anti-RBD and anti-S_WT antibody levels in the convalescent-vaccinated individuals. The distinct antibody patterns captured by our antigen screening biosensor indicate that the presence of these antibodies in combination with anti-M antibodies (anti-M_WT and anti-M_Om) can be used to detect and differentiate past COVID-19 infection and vaccination status.

### Comparison of antigen-specific antibody response patterns between clinically known samples and ML-classified blind samples

We used the antigen-specific anti-SARS-CoV-2 antibody measurements from the clinically known samples to train a supervised ML model (Random Forest, see Materials and Methods). We then measured 100 blind samples, about which we have no clinical information, and used the ML model to classify these blind samples into the four COVID-19 immunity groups. Fig. 4 (a) and (b) show the antigen-specific antibody levels for each group for the clinically known and blind samples, respectively. We applied a cosine similarity test among the corresponding groups (e.g., naïve vs. predicted naïve groups) to quantify the antibody level similarities between the clinically known and ML-classified cohorts. The cosine test scores were calculated as naïve=0.96, vaccinated=0.92, convalescent=0.92, and convalescent-vaccinated=0.99, where maximum and minimum dataset similarities can take values of 1 and 0, respectively. Moreover, in Fig. 4(c), we plot each antigen-specific antibody ‘s importance level for the sample classification as evaluated by the Random Forest algorithm. Based on the blind sample cohort measurements, the ML algorithm determined that anti-S_WT and anti-M_WT values are the most relevant in COVID-19 immunity status classification: 24% and 23%, respectively. This finding must be because of significantly high anti-S antibody values associated with vaccination and distinct anti-M peptide signals specifically marking past infections. Finally, the antibody response against the antigens for all individuals (known and blind with their immunity group predictions) is illustrated in Fig.4(d) for a better overview.

**Fig. 4.**
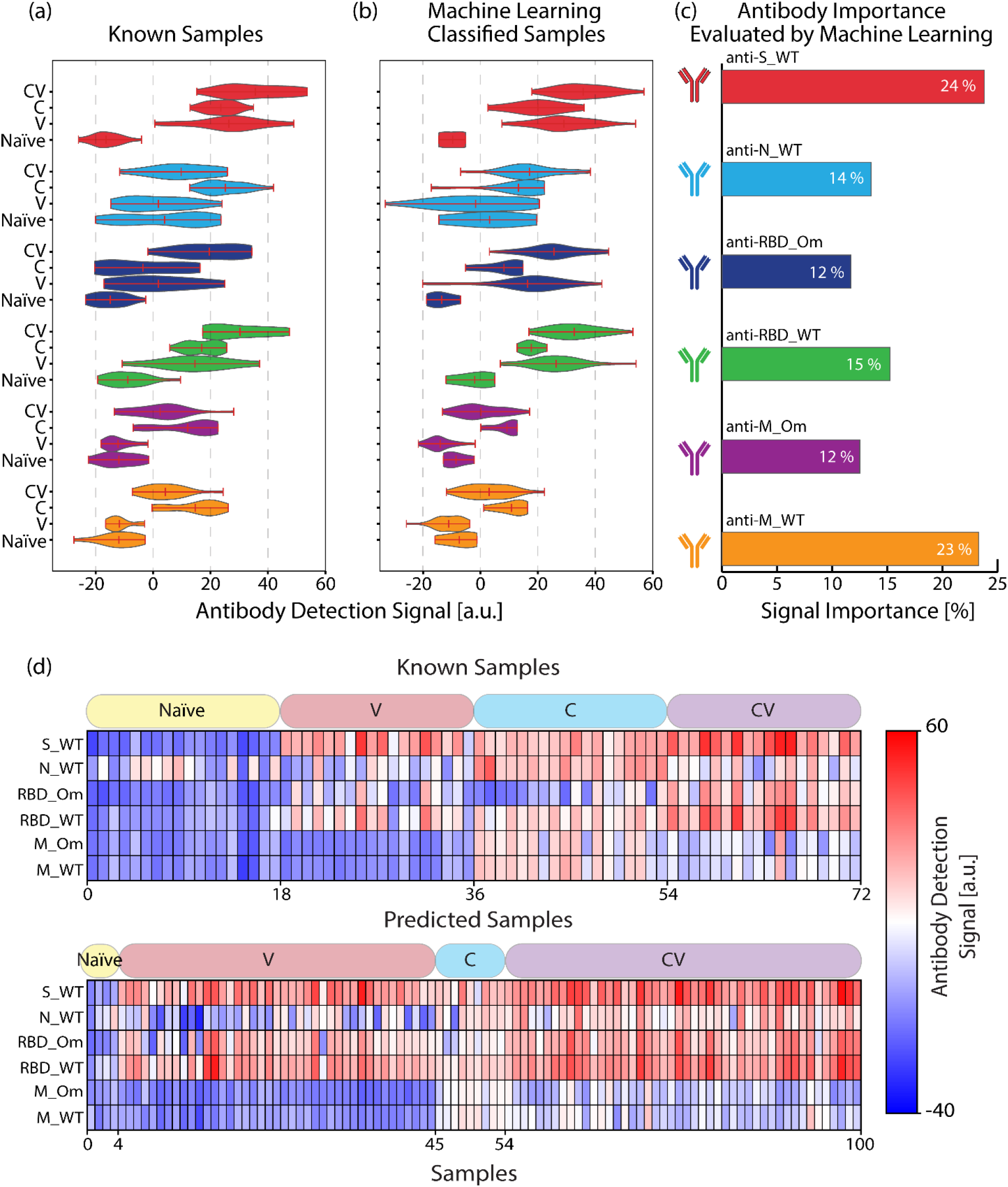
Comparison of antibody signals for **(a)** known and **(b)** ML-classified samples. **(c)** The importance of each antibody type for the ML classification model. **(d)** Heatmap represents the antibody response ‘s strength against SARS-CoV-2 antigens for known (n=72) and blind (n=100) samples. Each row represents an antibody concentration specific to an antigen, and each column is associated with a human serum or a plasma sample. The patient categories for both types of samples are indicated in the boxes above the heat maps.

### Comparison between our nanobiosensor ‘s COVID-19 immunity status prediction and official epidemiological data

According to publicly available epidemiological data, around 84% of the population in Dane County, WI, had at least one dose of vaccine by the end of our blind sample collection period (collection period: March 16, 2022-April 7, 2022) ^[37]^ (Fig 5 (a)). Furthermore, by using available epidemiological data for the period, it was estimated that 65% of the WI population had at least one COVID-19 infection ^[38–40]^ (Fig.5(b), for calculation of estimated cases, see Materials and Methods). Additionally, among all of the infections from the start of the pandemic until the end of the collection period, the Omicron variant ‘s share is estimated to be ∼30%, based on the calculations in which we projected the variant prevalence data in the US (Fig5(b), black curve) to the overall WI case numbers ^[41]^.

**Fig. 5.**
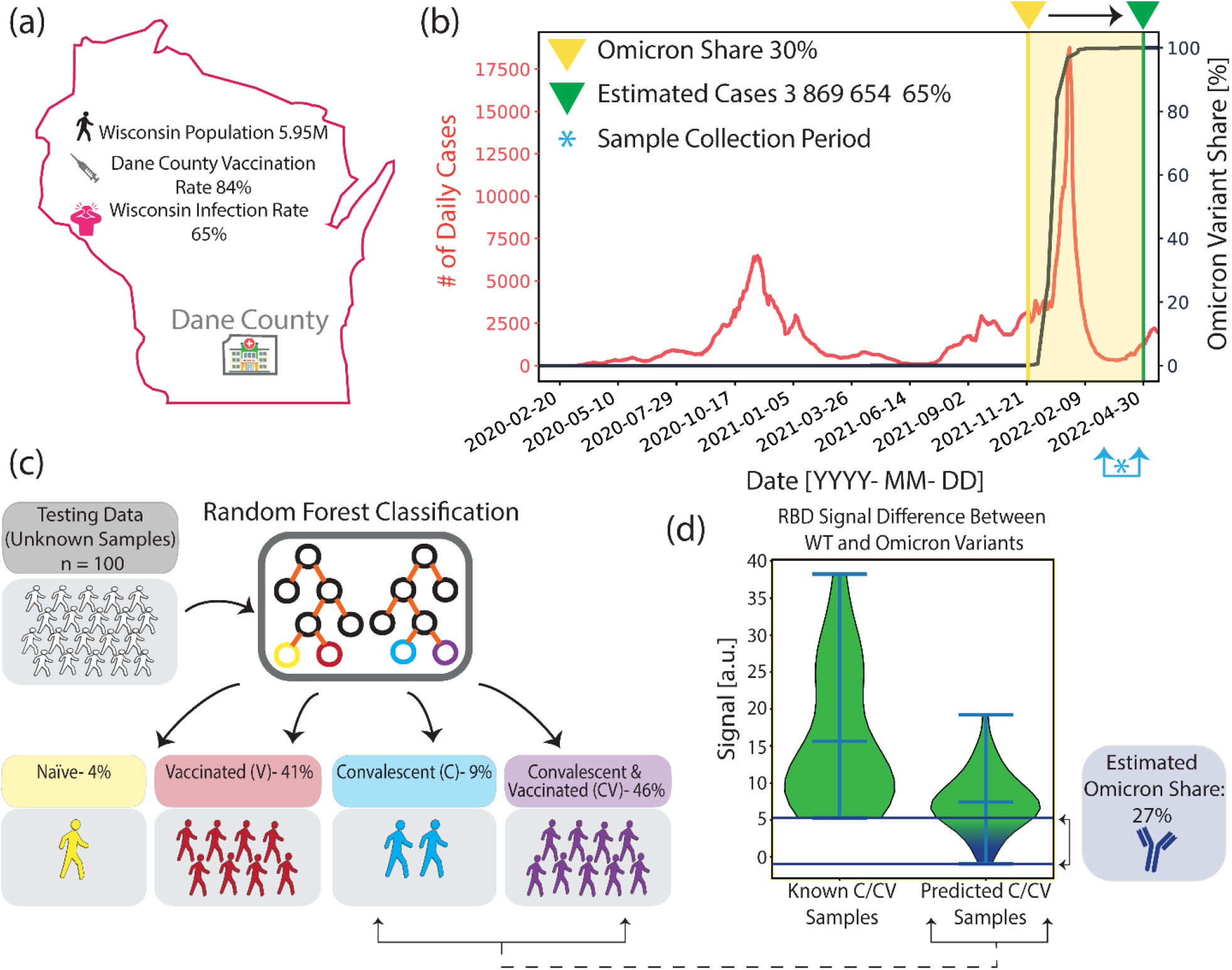
Comparison of past COVID-19 infection and vaccination rates between public records and our methods. **(a)** Infection and vaccination rates by the end of the sample collection period in Dane County, WI. **(b)** Wisconsin daily reported infection cases (red) and Omicron variant prevalence in the US (black) from the start of the pandemic until the end of the sample collection period. Asterisk indicates the sample collection period, between March 16, 2022 and April 7, 2022 **(c)** Summary of our COVID-19 immune profiling method findings based on 100 blind sample analyses. **(d)** Anti-RBD_WT and anti-RBD_Om antibody signal differences for samples with known past COVID-19 infections and ML-classified convalescent samples from the blind testing group. The Omicron infection rate among the blind sample group with past infection is estimated to be ∼27%.

Fig.5(c) depicts the COVID-19 immunity profiles of 100 blind samples, which were measured by our nanobiosensor and classified by the ML algorithm. We found that 4% of the blind samples are COVID-19 naïve, and 87% are vaccinated. Moreover, 55% of the samples had evidence of past infection, among which 46% had both past infection and vaccine, and 9% had only past infection but no vaccine. Our findings are very similar to the publicly available epidemiologic data (Table 1). Furthermore, we calculated the variant-dependent antibody response difference between anti-RBD_WT and anti-RBD_Om in the clinically known convalescent group, which doesn ‘t include any Omicron past infections because the sample collection was before the emergence of the Omicron variant. Subsequently, we calculated the same antibody response difference between anti-RBD_WT and anti-RBD_Om within the blind testing group that is predicted to have past infection. Using the minimum variant-based difference in the clinically known group as a threshold, we found that ∼27% of ML-classified samples with past infection fell below this threshold (Fig. 5(d)). Our Omicron infection estimate of ∼27% is similar to the ∼30% calculated for Dane County using the epidemiological data (Table 1).

**Table 1.** Summary of the COVID-19 past infection rate, vaccination rate, and Omicron variant share based on publicly available epidemiological data and our method.

| Data Type | Infection Rate [%] | Vaccination Rate [%] | Omicron Share [%] |
| --- | --- | --- | --- |
| Epidemiological COVID-19 Data | 65 | 84 | 30 |
| ML-Aided Nanobiosensor's Predictions | 55 | 87 | 27 |

## Discussion

One of the critical expectations from next-generation clinical biosensors is their contribution to the active decision-making process after processing and evaluating patient data rather than passively delivering measured results. However, for automated data evaluation tools, such as ML and artificial intelligence, to guide clinicians accurately, extensive datasets reflecting individuals ‘ detailed health status are needed. This important healthcare requirement highlights the critical role of diagnostic biochemical sensors that can simultaneously measure multiple biomarkers without requiring sophisticated instrumentation and large sample quantities. Notably, for equitable access to high-quality healthcare, such diagnostic devices need to be low-cost, portable, and robustly operatable outside of clinical laboratories.

To address this need, in this work, we demonstrate an innovative example of an ML-coupled accurate biosensor technology targeting a serodiagnostic application to profile COVID-19 immunity status at the population level. In total, we measured antibody levels of 172 different samples against six SARS-CoV-2 antigens, including peptides, protein domains, and whole proteins from the original WT and Omicron variant of SARS-CoV-2.

The clinically known samples in this study were part of an earlier study in which a subset of the antigen-specific COVID-19 antibody levels was measured by conventional ELISA ^[31]^. Our multiplexed nanobiosensor measurements generally concurred with the immunological signatures presented in the earlier clinical reports. For example, the presence of anti-M peptide antibodies after infection, the increase of anti-S antibodies after vaccination or infection, the decay of anti-N antibodies beyond 6 months after infection, and the vaccine-induced boost of anti-S antibodies in individuals who had COVID-19 infection were reproduced by our nanobiosensor findings.

In addition to replicating previously reported trends, we present variant-specific antibody detection sensitivity. For example, Fig. 3 shows that the antibody levels against RBD_WT were consistently higher than RBD_Om in response to both vaccination and infection. This outcome is expected as the measured samples were collected before the emergence of the Omicron variant. Also, our findings align with a recent work where variant-dependent antibody binding was shown using kinetic affinity measurements ^[42]^. Notably, this trend did not apply to the blind samples, some of which presented similar antibody levels against RBD_WT and RBD_Om (Fig. 5(d)). Since the blind samples were collected between March and April of 2022, right after the Omicron infection peak, the increase in antibody binding to the mutant RBD in the blind samples is likely due to a past Omicron infection. In contrast, we did not detect a statistically significant difference between antibody levels against M_WT and M_Om peptides, likely due to the difference of only one amino acid in their sequences. The sensitivity of our biosensor platform for different variants can be further tested in a future work, in which antibody concentrations against a specific antigen with a different number of mutations can be studied by measuring sera from individuals with known viral variant infections.

Moreover, incorporating an ML model has simplified the data evaluation process immensely. Our approach classifies each sample by comprehensively evaluating antibody patterns against all six antigens. This is in contrast to the conventional threshold-based classification approach, in which individual cutoff values are employed for each antigen-specific antibody measurement. Also, it is noteworthy that the ML algorithm detects anti-M_WT and anti-S_WT as the most defining features for classification. It would be interesting to see how the share of the importance of anti-RBD_Om will increase if populations with past Omicron infection or the now available bivalent vaccine had been part of the training data.

Finally, despite the relatively limited sample size (n=172) used in this work to derive epidemiological or medical conclusions, our approach shows strong potential for future studies with larger sample cohorts. Moreover, our pioneering approach of combining ML-based data evaluation tools with a powerful nanoplasmonic biosensor, which is label-free, rapid, and can simultaneously screen thousands of biomarkers, can significantly impact serodiagnostics at a broad global health scale beyond the COVID-19 pandemic. Combining our nanobiosensor with a portable optical reader ^[43]^ and a blood-filtering cartridge ^[26]^ could enable its applications at the point of care and support efforts for pandemic management and preparedness.

## Materials and Methods

### Nanobiosensor Fabrication

The Au nanohole arrays (Au-NHA) plasmonic devices, which consist of periodic nanoscale perforations into an Au thin film supported by a glass substrate, were used as nanobiosensors in this work. The Au-NHA nanoplasmonic device exhibits an extraordinary optical transmission (EOT), which is a sharp optical resonance mode. The EOT resonance mode can be excited by illuminating the Au-NHA sensor chip orthogonally, and the resonance peak can be measured via spectral analysis of the transmitted light. The resonance peak position of the Au-NHAs is extremely sensitive to the refractive index changes on the sensor surface, which supports the underlying biosensing mechanism used in this work. Details on the wafer-scale fabrication of the AuNHA chips can be found in ^[44]^. Briefly, 15 nm Titanium (Ti) and 100 nm Au were deposited sequentially on a 500 nm thick, 4-inch fused silica wafer (University Wafer, Massachusetts, USA) using a metal evaporator (in-house fabricated by Nano Fabrication Center UW Madison, Wisconsin, USA). The 15 nm-thick Ti layer functions as an adhesive layer for the Au layer on glass and suppresses the irrelevant surface modes induced by the SiO_2_-Au interface. The Au-NHAs (200 nm diameter and 600 nm period) were patterned using a 248 nm deep-ultraviolet stepper (ASML PAS 5500/300 DUV, Veldhoven, Netherlands). After the resist development, the Au-NHAs were transferred into the Ti/Au layer using an ion beam etching tool (Oxford Instruments PlasmaLab 300 IBE, Abingdon, UK). The wafers were then diced into 1×1 cm^2^ sensor chips by a dicing machine (Disco, Tokyo, Japan) using a resin blade. Before dicing, the wafers were coated with a photoresist layer for protection. Subsequently, the wafers were cleaned by a three-step cleaning process to ensure a clean sensor surface. First, the chips were immersed in MICROPOSIT™ remover 1165 (Rohm and Haas Electronic Materials, Massachusetts, USA) overnight. Second, the chips were treated in Oxygen plasma (250W, 1 minute, 80 sccm), and finally, the first step of the RCA cleaning was performed.

### Protein, Assay, Microarray Patterning, and Imaging

SARS-CoV-2 Nucleocapsid (N, Cat# 40588-V07E), Receptor Binding Domain Wild Type (RBD_WT, Cat# 40592-VNAH), Receptor Binding Domain Omicron (RBD_Om Cat# 40592-V08H121), and Spike (S_WT Cat# 40591-V08H) proteins (Sinobiological, Beijing, People ‘s Republic of China) were prepared by dilution from 1 mg/mL stock solution to 150 μg/mL with 1x phosphate buffer saline (PBS, for N), or Milli-Q Water (for RBD_WT, RBD_Om, and S_WT). All solutions contain 0.5% trehalose (m/v) and 0.005% Tween20 (v/v) to ensure even molecule adsorption in the microarray spots. The SARS-CoV-2 wild-type membrane peptide (8-M-16 sequence from ^[36]^, M_WT) with sequence ITVEELKKLLEQWNLV (Biomatik, Ontario, Canada) and SARS-CoV2 omicron membrane peptide (Q19E mutation, M_Om) with sequence ITVEELKKLLEEWNLV (Peptide 2.0, Virginia, USA) were both biotin conjugated. The M-protein peptide solutions were mixed with streptavidin (Sigma Aldrich, Missouri, USA) dissolved in 10mM acetate buffer with 25% trehalose (m/v) to achieve a final concentration of 200 μg/mL for each peptide. For a negative control measurement, rabbit anti-bovine IgG (Thermo Fisher Scientific, Massachusetts, USA), diluted to 200 μg/mL using milli-Q with 0.005% Tween20 (v/v) and 0.5% trehalose (m/v), was used in the microarrays. For a positive control measurement, protein A/G (Thermo Fisher Scientific, Massachusetts, USA) is diluted with 10mM acetate buffer and 0.5% glycerol with a final concentration of 500 μg/mL and spotted on the dedicated sensor areas. The microarray patterning of 200 pL droplets is performed using an iTWO-300 spotter (M2-Automation, Berlin, Germany) for all target proteins and controls. The droplet size on the chip is 150 μm diameter and spotted at 200 μm period. As shown in Fig. 1(b), each antigen was repeated five times in a microarray, which can be captured in a single image field of view (FOV). The positive and negative control proteins were spotted in a neighboring microarray and captured in a subsequent image. After the microarray patterning, chips were incubated for 2 hours to ensure the adsorption of the proteins on the Au sensor surface. To saturate streptavidin molecules with the biotin-conjugated M-protein peptides, we used a 1:4 molar concentration ratio between streptavidin and biotin-conjugated peptides. After microarray protein spotting, the sensor chips were incubated on a 12°C cooled substrate in a chamber with 55% humidity to prevent rapid evaporation of the droplets. After the protein incubation, the non-patterned areas of the sensor chips were blocked by immersion in 1% (v/v) bovine serum albumin (BSA) in 1x PBS solution for 20 minutes. Subsequently, the chips are washed in 1x PBS with 1% (v/v) Tween20 solution for 5 minutes under constant agitation to remove excess proteins. Next, the patterned chips were optically characterized to collect reference images, incubated with the serum sample for 30 minutes, washed with 1x PBS under agitation for 5 minutes, and imaged again.

### Optical Setup

A broadband laser SuperK FIANIUM15 (NKT Photonics, Birkerød, Denmark) coupled to an LLTF filter (Photon etc., Montreal, Canada) was used as a narrowband light source with a full-width half maximum (FWHM) of ∼ 0.25 nm. The imaging is performed at λ=650 nm, which is located near the plasmonic resonance peak of the Au-NHAs. The nanobiosensors were illuminated via a beam collimator and imaged in transmission mode with a 10x magnification objective on the optical light path of a TE-200 inverted microscope (Nikon, Tokyo, Japan) using a Prime BSI Express CMOS camera (Teledyne Photometrics, Arizona, USA). The camera exposure time is set to 25 ms, and the acquisition was made using a custom-made MATLAB (MathWorks, Massachusetts, USA) user interface, where the camera and filter functionalities were integrated into a single application using the app designer feature. For intensity normalization of the acquired images, the source is imaged through a neutral density (ND) filter of optical density (OD) 3 without the sensor chip.

### Data Processing

The image data processing was performed using Python libraries in Jupyter Notebook similar to the single wavelength approach of previous work ^[26]^ with minor modification. First, the image pixel values of the sensors were normalized to the source. Second, spatially aligned sensor images collected before and after incubation with the sample were subtracted from each other. Third, for each antibody measurement, the resulting intensity difference was sampled from 5 different circular areas with a radius of 25 pixels from each microarray spot. These pixel-wise intensity differences were then subtracted by the average intensity difference from the unpatterned sensor areas within the same image, which corresponds to the non-specific background noise. Subsequently, the values that lie between 2σ and 3σ of the distribution of the subtracted data were selected and averaged. This average value is our antibody signal against one protein and was multiplied with -100.

### Statistical Analysis

To examine the difference of the antibody signals between proteins, peptides, protein domains and groups, p-values were calculated using one-way ANOVA using MATLAB (MathWorks, Massachusetts, USA) software.

### Machine-Learning Model

For the machine-learning model, Random Forest Classifier from Scikit-Learn with Python in Jupyter Notebook was used with the training data to create an automatic predictive model. All the parameters were left at default, and the number of trees used was 10,000 after confirming the convergence of the Out of Bag (OOB) score of 90% at 1,000 trees. The training data used were six antigen-specific antibody values measured from n=72 different serum samples with clinically labeled COVID-19 immunity status as in the following four groups: convalescent (n=18), convalescent-vaccinated (n=18), naïve (n=18), and vaccinated (n=18). The model then predicts the COVID-19 immunity status for 100 blind serum samples (those without information on their COVID-19 immunity status). The result of the prediction is then validated with published epidemiological data and is discussed in the results and discussion sections.

The number of estimated COVID-19 cases by the end of the sample collection period in WI is taken from the CDC ^[39]^. However, the CDC ‘s prediction covers only until the end of February 2022 (3,854,000 cases). Therefore, the number was extrapolated to two weeks before the end of sample collection (March 14^th^, 2022) as follows: by the end of February 2022, the cumulative number of reported cases in WI is 1,571,977 ^[39]^, whereas by March 14^th^ 2022, the number grew to 1,578,362 ^[38]^. The ratio between these two numbers is calculated to be 1.004 and is multiplied with the aforementioned CDC ‘s prediction to obtain the extrapolated number of 3,869,654 cases. This comprises around 65% of Wisconsin ‘s population at the time ^[40]^. Two weeks is deducted from the end of sample collection to take into account that it takes on average about 2 weeks to generate antibodies from time of infection. The number of vaccination rate in Dane county is taken from ^[37]^. For the total number of Omicron cases, the weekly reported cases in Wisconsin (^[38]^, see Fig.5 (b), red curve) from the beginning of the pandemic until the end of the sample collection period is multiplied with the prevalence of Omicron as a function of time (^[41]^, see Fig.5 (b), black curve). The ratio of the total reported Omicron case number to the total case number is then calculated to be about 30%.

### Clinical Samples

Human studies were performed according to the Declaration of Helsinki and were approved by the University of Wisconsin Institutional Review Board. COVID-19 naïve samples were collected prior to the pandemic and were obtained from the UWCCC TSB. COVID-19 convalescent samples were obtained from the UW COVID-19 Convalescent Biobank, which has been previously described ^[7,31]^, via the UWCCC TSB. In brief, all subjects with COVID-19 had a positive SARS-CoV-2 PCR test in the spring of 2020. The convalescent sera were collected at ∼1 or ∼3 months post-symptom resolution. The convalescent vaccinated samples were collected 12 months post-COVID-19 symptom resolution, and subjects had received at least one vaccine dose ≥ 5 days before sample collection. Samples from individuals vaccinated against SARS-CoV-2 (>3 weeks after two mRNA vaccine doses or one Ad26.COV2.S dose) and without known COVID-19 were collected in the summer of 2021 as described in ^[31]^ and obtained from the UW COVID-19 Convalescent Biobank. Blind samples were excess serum and plasma being discarded after cholesterol tests were performed at UW Health (Dane County, WI) and were collected between March 16 and April 7, 2022.

## Data Availability

All data are available in the main text and Supplementary 
Material.

## Acknowledgments

The authors thank the UCSB Nanofabrication Facility, an open-access laboratory, specifically Demis D. John, Ph.D., and Biljana Stamenic for performing the lithography steps of the nano biosensors. Additionally, we thank Naomi Buckli, Don Dorn, and Janis Parkinson at UW Health Clinical Laboratories for their assistance with collecting the blind samples and the Translational Science Biocore (TSB) BioBank of the University of Wisconsin Carbone Cancer Center for providing specimens and associated clinical data. Moreover, we thank Claude Dufresne, Ph.D. from Axivend, for assistance in operating the microarray dispenser. Finally, we thank Prof. Eduardo R. Arvelo from the Electrical & Computer Engineering Department at the UW-Madison for assistance in illustration and guidance in machine learning methods.

## Author contributions

Conceptualization: AB, WA, MAS, FY

Methodology: AB, WA, MAS, FY

Investigation: AB, WA, MV, KBG, SJB

Visualization: AB, WA

Supervision: MAS, FY

Writing—original draft: AB, WA, FY

Writing—review & editing: AB, WA, SJB, MAS, FY

## Competing interests

M. A. S. is listed as an inventor on a patent filed related to this study (PCT/US2021/051143; IDENTIFICATION OF SARS-COV-2 EPITOPES DISCRIMINATING COVID-19 INFECTION FROM CONTROL AND METHODS OF USE). All other authors declare that they have no competing interests.

### Data and materials availability

All data are available in the main text and Supplementary Material.

